# Effectiveness of BBIBP-CorV, BNT162b2 and mRNA-1273 vaccines against hospitalisations among children and adolescents during the Omicron outbreak in Argentina

**DOI:** 10.1101/2022.04.18.22273978

**Authors:** Soledad González, Santiago Olszevicki, Alejandra Gaiano, Ana Nina Varela Baino, Lorena Regairaz, Martín Salazar, Santiago Pesci, Lupe Marín, Verónica V. González Martínez, Teresa Varela, Leticia Ceriani, Enio Garcia, Nicolás Kreplak, Alexia Navarro, Elisa Estenssoro, Franco Marsico

## Abstract

**Background:** Although paediatric clinical presentations of COVID-19 are usually less severe than in adults, serious illness and death have occurred. Many countries started the vaccination rollout of children in 2021; still, information about effectiveness in the real-world setting is scarce. The aim of our study was to evaluate vaccine effectiveness (VE) against COVID-19-associated-hospitalisations in the 3-17-year population during the Omicron outbreak.

**Methods:** We conducted a retrospective cohort study including individuals aged 3-17 registered in the online vaccination system of the Buenos Aires Province, Argentina. mRNA-1273 and BNT162b2 were administered to 12-17-year subjects; and BBIBP-CorV to 3-11- year subjects. Vaccinated group had received a two-dose scheme by 12/1/2021. Unvaccinated group did not receive any COVID-19 vaccine between 12/14/2021-3/9/2022, which was the entire monitoring period. Vaccine effectiveness (VE) against COVID-19-associated hospitalisations was calculated as (1-OR) x100.

**Findings:** By 12/1/2021, 1,536,435 individuals aged 3-17 who had received zero or two doses of SARS-CoV-2 vaccines were included in this study. Of the latter, 1,440,389 were vaccinated and 96,046 not vaccinated. VE were 78·0% [68·7-84·2], 76·4%[62·9-84·5] and 80·0%[64·3-88·0] for the entire cohort, 3-11 subgroup and 12-17 subgroup, respectively. VE for the entire population was 82·7% during the period of Delta and Omicron overlapping circulation and decreased to 67·7% when Omicron was the only variant present.

**Interpretation:** This report provides evidence of high vaccine protection against associated-hospitalisations in the paediatric population during the Omicron outbreak but suggests a decrease of protection when Omicron became predominant. Application of a booster dose in children aged 3-11 warrants further consideration.

**Research in context:** *Evidence before this study:* There is limited evidence on the effectiveness of vaccines in the pediatric population, particularly in children aged 3-11 years after the SARS-CoV-2 B.1.1.529 (Omicron) variant’s emergence. We searched preprint and peer-reviewed published articles in PubMed, medRxiv, and SSRN for observational studies, with no language restrictions, using the term “COVID-19 OR SARS-CoV-2” AND “vaccine effectiveness” OR “vaccine impact” AND “children” OR “pediatric” AND “Omicron” published between December 1, 2021, and April 1, 2022. We found 4 studies that included subjects in the 3-17-year population who received a two-dose-scheme of any of the available vaccines-according to each country’s authorisation. Three studies were from the US; two were test-negative-case-control studies and one was a retrospective non-peer-reviewed cohort study. The reported vaccine effectiveness (VE) for 2-doses of BNT162b2-mRNA (Pfizer-BioNTech) in preventing hospitalisations during Omicron predominance was 48-78%; and it was 40-92% for 5-11 and 12-17-year subgroups, respectively. The fourth was a cohort study still in preprint form conducted in Chile and utilized an inactivated vaccine, CoronaVac (SinoVac), widely used in Latin-America. VE for two doses of CoronaVac in the 3-5-year subgroup against hospitalisations was 64% and 69% against ICU admissions.

*Added value of this study:* Up to date, there are no published studies about the effectiveness of the BBIBP-CorV vaccine against hospitalisation in the pediatric population. Additionally, there are no real-world studies from low and middle-income countries about VE in the 12-17 aged population during the Omicron outbreak. This study shows that VE after 14 days or more from two-dose-scheme was 78·0% [68·7-84·2], 76·4% [62·9-84·5] and 80·0% [64·3-88·0] for the 3-17-year entire group, and for 3-11-year (BBIBP-CorV) and 12-17-year (mRNA vaccines) subgroups, respectively. VE for the 3-17-year entire group was 82·7% during the period of Delta and Omicron overlapping circulation and decreased to 67·7% when Omicron was the only variant present. These effects were consistent across all subgroups.

*Implications of all the available evidence:* Our results provide evidence of high vaccine protection against COVID-19 associated-hospitalisations in the pediatric population during the Omicron outbreak, but suggest a decrease of protection when Omicron became predominant. Application of a booster dose in children aged 3-11 warrants further consideration.

## INTRODUCTION

Vaccines against SARS-CoV-2 have proven to be a critical tool for controlling the COVID-19 pandemic. Studies of vaccine efficacy, safety and real-world effectiveness have been published.^1^ However, the emergence of the highly transmissible Omicron (B.1.1.529) variant of concern (VOC), which has been able to partially avoid the immune response achieved after vaccination or natural infection, has caused an increase of COVID-19 cases worldwide.^2,3^ Recent studies have shown that vaccine effectiveness (VE) could be lower against Omicron.^4-6^ Omicron was isolated in Argentina in the last week of November 2021; and from the third week of December, it has rapidly gained predominance.^7^

Although paediatric clinical presentations of COVID-19 are usually less severe than in adults, serious illness and the Multisystem Inflammatory Syndrome (MIS-C) have occurred in children after the primary infection.^8^ In addition, during the last COVID-19 wave, the rate of COVID-19-associated hospitalisations in children in the United States, South Africa and Hong Kong is notably higher in comparison to hospitalisations occurring in the Delta period.^9–12^

Following publication of phase 2 and 3 studies demonstrating the safety and efficacy of the different vaccines, the Emergency Use Authorization (EUA) for the 3-17 age groups was approved by different international health organisations.^13–17^

Argentina started vaccination rollout in the adolescent subpopulation in August 2021 with mRNA-1273 (Spikevax) from Moderna and BNT162b2 (Comirnaty) from Pfizer-BioNTech. Argentina was also one of the first countries to introduce paediatric COVID-19 vaccination in the group of 3-11-year-old children in October 2021 with BBIBP-CorV (Sinopharm) from the Beijing Institute of Biological Products.^18^ As of March 9 2022, 1,487,859 children and 1,279,170 adolescents have been fully vaccinated in the Province of Buenos Aires, which is equivalent to 57% and 80% of the 3-11-year and 12-17-year of the total population, respectively.

Information about vaccine effectiveness from real-world studies of the paediatric population is scarce, especially in children younger than 12.^19–23^ Additionally, data on real-world effectiveness of BBIBP-CorV, an inactivated vaccine, is not available in the paediatric population. Recently, four studies about vaccine effectiveness of BNT162b2 (Pfizer-BioNTech) and Coronavac (Sinovac) carried out in 3-11-year-old children during the Omicron period demonstrated that COVID-19 vaccines elicited less protection in the younger group and exhibited faster waning.^24–27^

The aim of this study is to evaluate the effectiveness of COVID-19 vaccines authorised in Argentina against hospitalisations in 3-17-year-old children and adolescents in Buenos Aires Province during the Omicron outbreak. Our hypothesis is that the two-dose-scheme of vaccination in this age group is associated with lower incidence of hospitalisations.

## METHODS

### Study Design and Participants

This is a retrospective cohort study aimed at determining the effectiveness of the COVID-19 vaccines against COVID-19-associated-hospitalisations in children and adolescents in the Province of Buenos Aires, Argentina, the most densely populated area of the country, with 17·7 million inhabitants.

To address the vaccination campaign against COVID-19, the Ministry of Health developed its own registration system (Vacunate PBA). Registration is voluntary and can be carried out via Android and IOS applications, or through a specially designed website. To receive a vaccine, province residents need to register their age, gender, occupation, and underlying conditions.

Vaccine information reported in the Vacunate PBA included date of vaccination, number of doses, type of vaccine, vaccine lot number, and vaccination centre.

Additional information about COVID-19 confirmed cases (by laboratory testing, epidemiological and/or clinical criteria), hospitalisations, and deaths of subjects included in the present study was obtained from the National System of Health Surveillance, up to March 9, 2022. Further data was added by the Bed Management System, a province-level monitoring system for hospital admission, discharge and death.

Following the prioritised and sequentially progressive COVID-19 vaccination campaign, Argentina began immunisation of adolescents in August 2021. mRNA vaccines (mRNA-1273 from Moderna and BNT162b2 from Pfizer-BioNTech) were applied. Three to eleven-year-old children began vaccination with BBIBP-CorV, an inactivated vaccine, on 13 October 2021.^18^ The primary vaccination scheme consisted of 2 doses with a 21- or 28-day interval in immunocompetent subjects, and 3 doses with a 28-day interval in immunocompromised children.

Inclusion criteria for the study were: age between 3 and 17 years, registration in Vacunate PBA by 1 December 2021 to receive COVID-19 vaccine, and residence in the Province of Buenos Aires. The vaccinated group had to have the vaccination two-dose scheme by 1 December 2021; and should not have received a third or a booster dose. The unvaccinated group did not receive any COVID-19 vaccine during their monitoring period. Subjects receiving a different vaccine from the one corresponding to their age group were excluded.

The monitoring period started on 14 December 2021 and ended 14 days after the date of the positive SARS-CoV-2 laboratory test, or on the date of the case confirmation by clinical and/or epidemiological criteria. The date of confirmed-SARS-CoV-2 infections was identified by the symptom-onset date or, if not available, the date of the sample collected for COVID-19 test. For subjects with no positive diagnosis for COVID-19, monitoring ended on 9 March 2022, the last day of the database update.

The primary outcome measure was the proportion of the study participants with COVID-19-associated hospitalisation occurring within 14 days of confirmed SARS-CoV-2. Because of the low rate of hospitalisations in the paediatric population, any admission to moderate or intensive care units were included as a single outcome.

Finally, we assessed vaccine effectiveness (VE) during a period coinciding with overlapping circulation of B.1.617.2 (Delta) and B.1.1.529 (Omicron) (14 December 2021–18 January 2022) and Omicron-predominant period (19 January–9 March 2022). The National Ministry of Health’s genomic surveillance strategy has focused on detecting VOCs through laboratory-confirmed SARS-CoV-2 infections by RT-PCR selected for regular surveillance of circulating variants in the general population, serious or unusual clinical presentations, vaccinated individuals, suspected cases of reinfection and travellers.^7^

#### Vaccine authorisation

BBIBP-CorV of Beijing Institute of Biological Products was authorised for emergency utilisation in adults by the National Ministry of Health in February 2021 (report number 688/2021) after recommendation of the National Administration for Drugs, Food and Technology. On 1 October 2021 the authorisation was extended for the use in children older than 3 years.

mRNA-1273 (Spikevax) from Moderna and BNT162b2 (Comirnaty) from Pfizer-BioNTech were authorised for the use in adolescents (12-17 years) from 23 July and 28 May 2021 respectively by the Committee for Medicinal Products for Human Use (CHMP) from European Medicines Agency.

#### Ethics

The Central Ethics Committee of the Ministry of Health of the Province of Buenos Aires evaluated and approved the protocol of the present study on 18 March 2022. The report number ACTA-2022-07714920-GDEBA-CECMSALGP.

##### Informed Consent

This study was exempted of informed consent due to its retrospective nature, and given it is a public health-related official programme.

##### Anonymization of data

Data were anonymized by the following procedure: The personal ID number was used to link the databases of follow-up and vaccination. After this process, we removed the personal ID number and created an ID reference number for each individual. This reference number is not associated with any personal information.

### Statistical analysis

Age subgroup, gender, previously registered SARS-CoV-2 infections and presence of comorbid conditions were compared between vaccinated and unvaccinated groups by means of Chi squared or Student’s t-test for independent samples, as appropriate. Age subgroup, gender, and presence of comorbid conditions were used as covariates in adjusted logistic regression models.

A p-value < 0·05 was considered significant.

Vaccine effectiveness was calculated as: (1 –OR) x 100. For the main analysis, we fitted logistic regression models to obtain unadjusted and adjusted odds ratios (OR) and their respective CI_95%_ for COVID-19-associated hospitalisations between vaccinated and unvaccinated subjects. Different adjustments were assessed using the Akaike Information Criterion (AIC). The final adjusted model used age (3-11 and 12-17 age subgroups), comorbid conditions, and gender as binary covariates. Further sub-analyses were assessed by stratifying the cohort by each binary variable. Another sub-analysis excluding those subjects with registered SARS-CoV-2 previous infection was performed. Differences in OR for hospitalisation between subgroups (3-11 years old against 12-17 years old, males against females, subjects with against subjects without comorbidities) were evaluated statistically with pairwise comparisons. Assuming normal distribution of the natural logarithm (ln) of the OR, Student’s t-tests for the difference between the in (OR) were conducted.

Data preprocessing was carried out with PostgreSQL (Portions Copyright © 1996-2022, The PostgreSQL Global Development Group). All statistical analyses were performed with R (R Development Core Team, 3.6.1 version) software. Open source code for performing statistical analysis is available on github (https://github.com/MarsicoFL/Vactools).

### Funding

This study did not receive any fundings.

### Data sharing statement

The confidentiality of the data obtained through the VacunatePBA and Bed Management System records was guaranteed. The use of the data was exclusively for the purposes of this research, preserving the anonymity of the persons included. Data will be available for researchers who provide a methodologically sound proposal after it is approved. The data that could be shared is Individual participant data that underlie the results reported in this article, after de-identification (text, tables, figures, and appendices). Proposals should be directed to; to gain access, data requestors will need to sign a data access agreement.

### Declaration of Competing Interest

NK, LC, TV, AG and SP declared being involved in the decision making process of the vaccination campaign in the Province of Buenos Aires, Argentina. All other authors report no competing interests.

## RESULTS

By 1 December 2021, 3,093,932 subjects aged 3-17 years registered in Vacunate PBA; 1,536,435 who had received zero or two doses of SARS -CoV-2 vaccines were included in this study. Of the latter, 1,440,389 were vaccinated and 96,046 not vaccinated, providing a 15:1 ratio. The flowchart of the study is shown in Figure 1.

**Figure 1:**
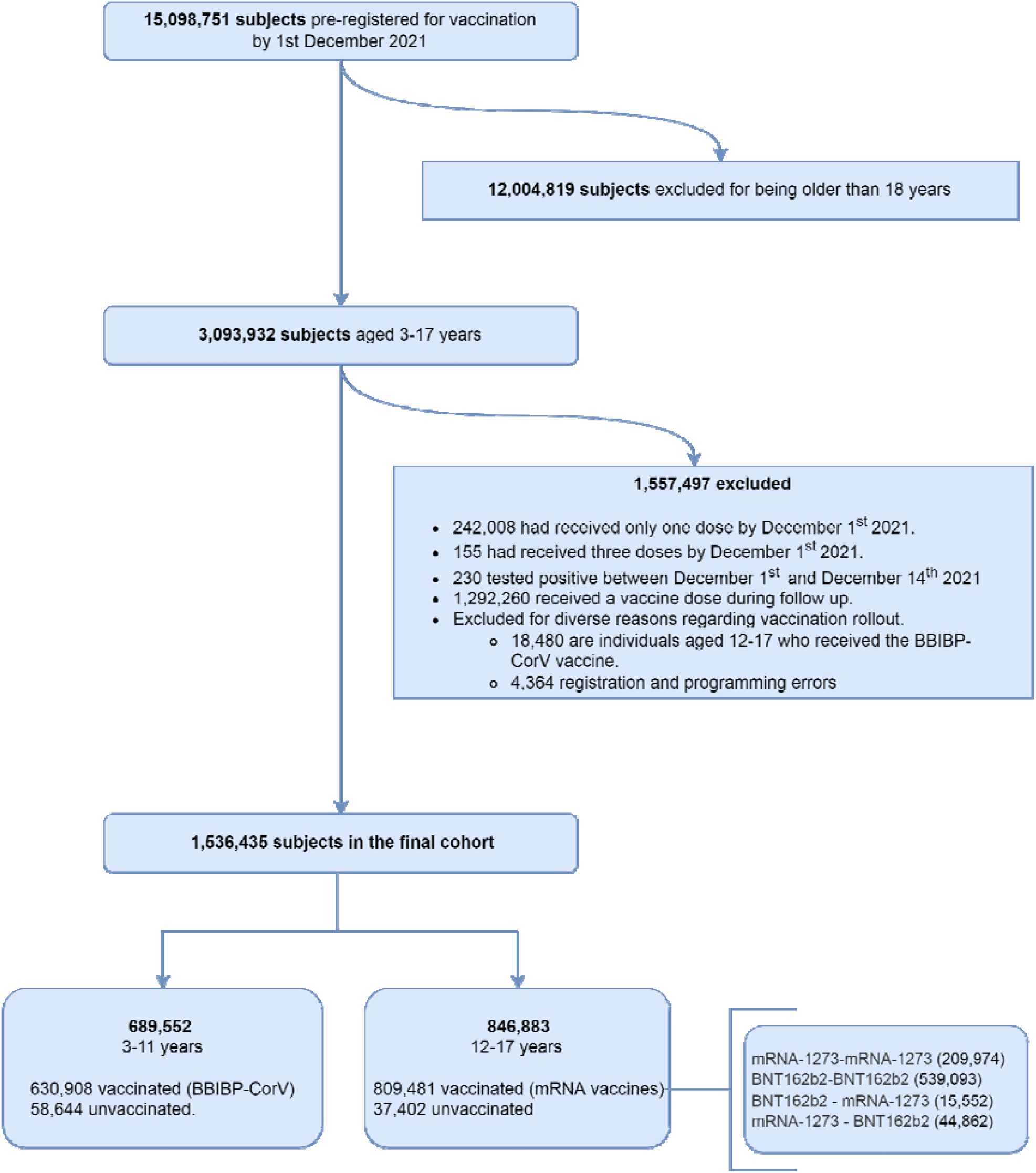
Flowchart of the study

The increasing prevalence of Omicron together with the number of infections and hospitalisations by SARS-CoV-2 over the study period are shown in Figure 2 a-c. Of note, since epidemiological week 4 there was no circulation of Delta VOC (B.1.617.2). Regarding the Omicron sub-lineage, BA.1 and BA.1.1 prevailed, while BA.2 represented less than 0·5% of the samples.^12^

**Figure 2:**
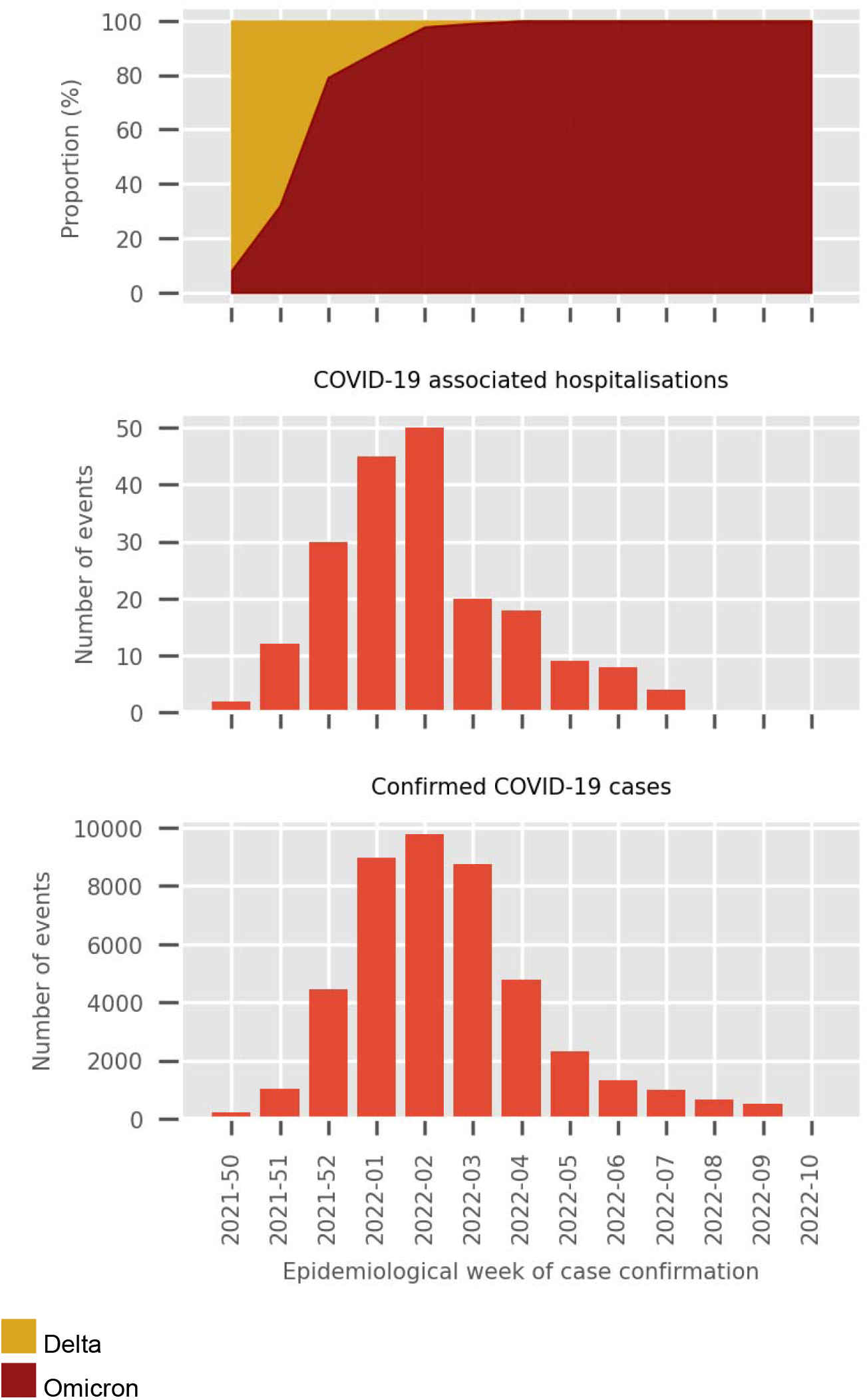
Characteristics of the COVID-19 pandemic in Argentina and the Province of Buenos Aires.

Characteristics of the entire group and comparisons between vaccinated and unvaccinated subjects are shown in Table 1. The vaccinated cohort was slightly older (11·7 years old vs 9·7, p < 0·001), had a lower proportion of males (49·8% vs 53·6%, p < 0·001), a higher percentage of subjects with comorbid conditions (17·8% vs 10·0%, p < 0·001) and a lower proportion of subjects with previously registered SARS-CoV-2 infection (0·1 vs 0·1%, p = 0·031) than the unvaccinated.

**Table 1.** Epidemiological characteristics of the entire population and of the vaccinated and unvaccinated age subgroups.

|  | Overall population |  |  |  | 3-11 years |  |  |  | 12-17 years |  |  |  | p-value |
| --- | --- | --- | --- | --- | --- | --- | --- | --- | --- | --- | --- | --- | --- |
|  | Overall<br>(n =<br>1,536,435) | Unvaccinated<br>(n =<br>96,046) | Vaccinated<br>(n =<br>1,440,389) | p-value | Overall<br>(n =<br>689,552) | Unvaccinated<br>(n =<br>58,644) | Vaccinated<br>(n =<br>630,908) | p-value | Overall<br>(n =<br>846,883) | Unvaccinated<br>(n =<br>37,402) | Vaccinated<br>(n =<br>809,481) | p-value |  |
| Age in years | 11.6 (4.1) | 9.7 (4.6) | 11.7 (4.0) | <0.001 | 7.6 (2.4) | 6.5 (2.6) | 7.7 (2.4) | <0.001 | 14.8 (1.6) | 14.7 (1.7) | 14.8 (1.6) | <0.001 | <0.001 |
| Male (%) | 769170 (50.1) | 51459 (53.6) | 717711 (49.8) | <0.001 | 348857 (50.6) | 31129 (53.1) | 317728 (50.4) | <0.001 | 420313 (49.6) | 20330 (54.4) | 399983 (49.4) | <0.001 | <0.001 |
| Comorbidities (%) | 266097 (17.3) | 9590 (10.0) | 256507 (17.8) | <0.001 | 73162 (10.6) | 5026 (8.6) | 68136 (10.8) | <0.001 | 192935 (22.8) | 4564 (12.2) | 188371 (23.3) | <0.001 | <0.001 |
| Previously infected (%) | 1589 (0.10) | 120 (0.12) | 1469 (0.10) | 0.031 | 228 (0.03) | 39 (0.07) | 189 (0.03) | <0.001 | 1361 (0.16) | 81 (0.22) | 1280 (0.16) | <0.001 | <0.001 |

In the 3-11 age subgroup, 630,908 (91·5%) were vaccinated and 58,644 (8·5%) remained unvaccinated. The vaccinated children were older (7·7 vs 6·5, p < 0·001), more frequently male and had a higher incidence of comorbidities. The proportion of previously infected children was similar between those vaccinated and unvaccinated.

With respect to the 12-17 age subgroup, 809,481 (95·6%) subjects were vaccinated and 37,402 (4·4%) remained unvaccinated. The vaccinated group was slightly older and predominantly female. Comorbid conditions were remarkably more frequent compared to the unvaccinated (23·3 vs 12·2 p < 0·001). The proportion of previously infected subjects was lower in the vaccinated group.

Days from vaccination to the end of follow-up period were 114 (±17) for the entire group, and 110 (±19) and 120 (±26) for the 3-11 and 12-17 age subgroups, respectively.

There were 43,787 COVID-19 confirmed cases out of 1,536,435 children and adolescents registered in Vacunate PBA (2·8%). Of these, 3,335 out of 96,046 (3·4%) were unvaccinated and 40,452 out of 1,440,389 (2·8%) were vaccinated.

Regarding hospitalisation, there were 44 hospitalisations in the infected unvaccinated subgroup (1·32%) and 154 in the infected vaccinated subgroup (0·38%). One child (0·03%) was admitted to the ICU in the infected unvaccinated subgroup and 6 children (0·01%) in the infected vaccinated subgroup. The only death registered occurred in the unvaccinated subgroup.

Cumulative-incidence curves of COVID-19-associated hospitalisation for the entire cohort and according to age subgroups are illustrated in figure 3a and 3b, respectively.

**Figure 3a:**
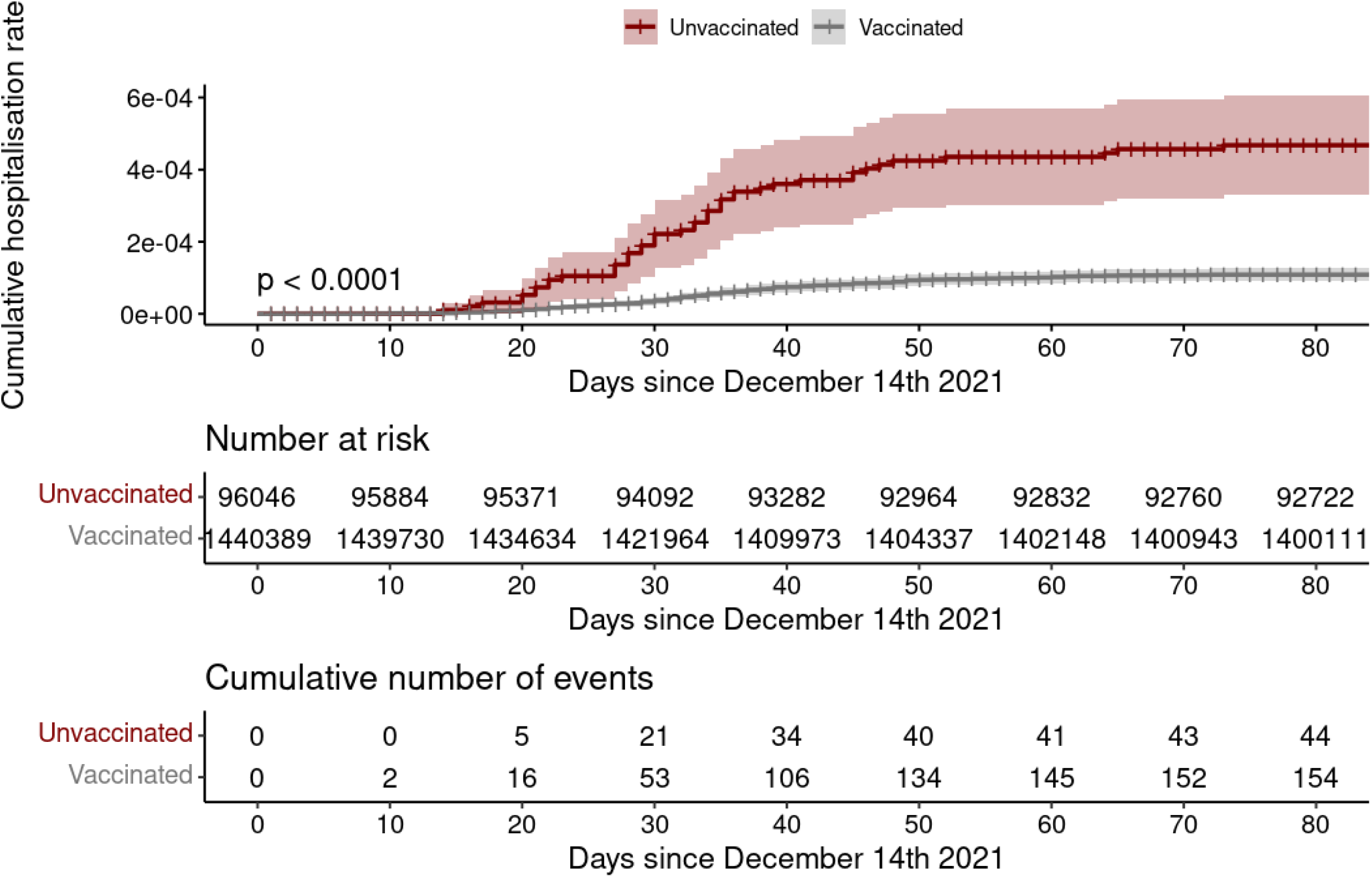
Cumulative incidence of hospitalisations by vaccination status.

**Figure 3b:**
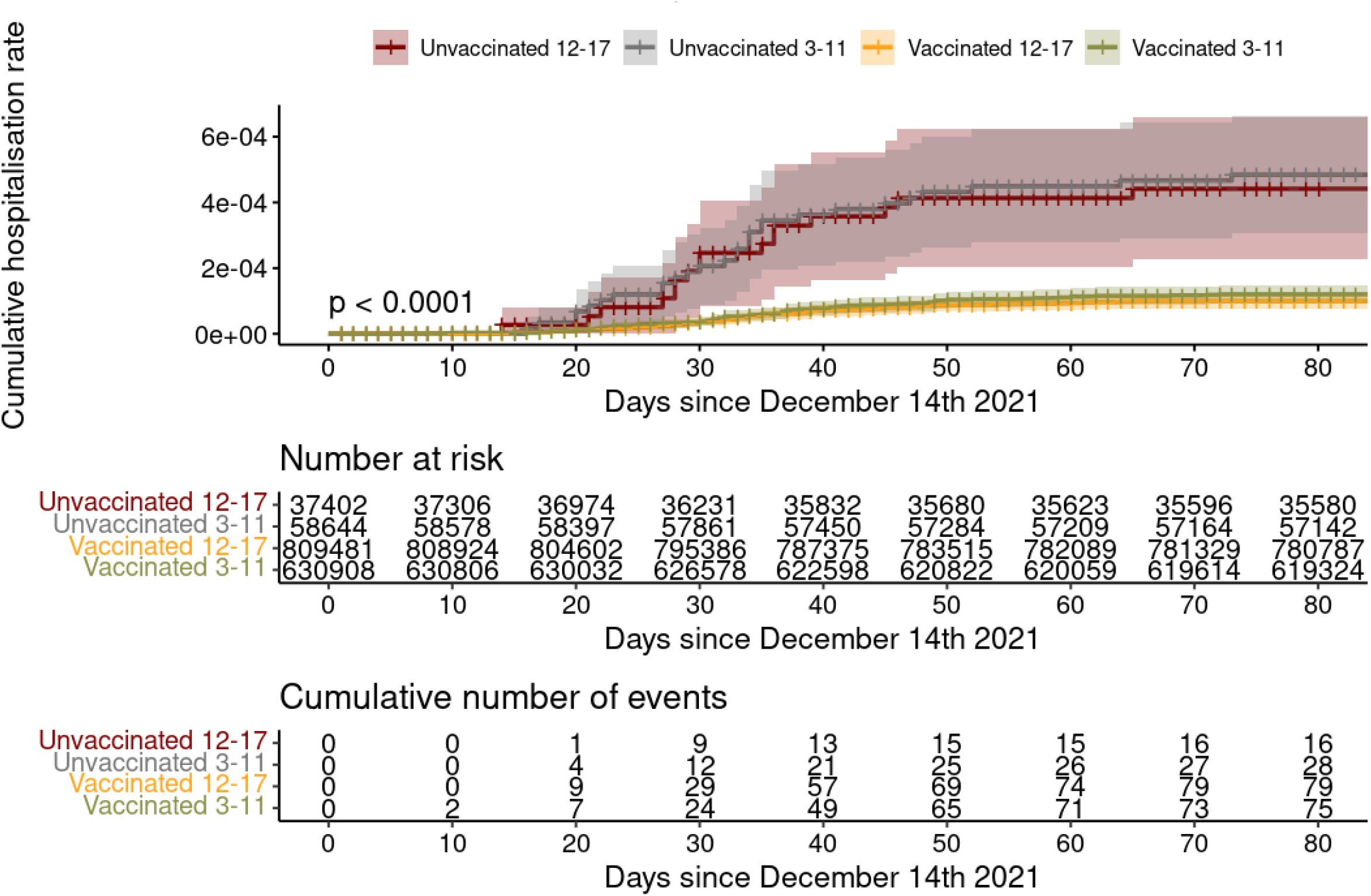
Cumulative incidence of hospitalisations by vaccination status in each age subgroup.

We found that the effectiveness of the vaccination against COVID-19-associated hospitalisations adjusted for age group, gender and presence of comorbid conditions was 78·0% [68·7-84·2]. These values, together with crude, are shown in Table 2.

**Table 2.** Analysis of vaccine effectiveness against hospitalisations in vaccinated and unvaccinated subjects

|  | Events | Crude effectiveness | Adjusted effectiveness * |
| --- | --- | --- | --- |
| Unvaccinated (n=96,046) | 44 | 76.6<br>[67.3-83.3] | 78.0<br>[68.7-84.2] |
| Vaccinated (n=1,440,389) | 154 |  |  |

Events are expressed as numbers.
Effectiveness is expressed as % [CI<sub>95%</sub>]
\*Adjusted for age group, gender, and presence of comorbid conditions.

Vaccination was effective against hospitalisations across all subgroups analysed.

Effectiveness for the 3-11 age subgroup was 76·4% [62·9-84·5] and 80·0% [64·3-88·0] for the 12-17 age subgroup. Crude and adjusted VE for subgroups of age, gender, and presence or not of comorbidities are shown in Table 3 and Figure 4.

**Table 3.** Vaccine effectiveness against COVID-19 associated hospitalisations for each subgroup of age, gender and comorbidities.

| Subgroup |  | Events | Crude effectiveness | Adjusted effectiveness* |
| --- | --- | --- | --- | --- |
| 3-11<br>(n = 689,552) | Unvaccinated<br>(n =58,644) | 28 | 75.1 [61.0-83.7] | 76.4 [62.9-84.5] |
|  | Vaccinated<br>(n =630,908) | 75 |  |  |
| 12-17<br>(n = 846,883) | Unvaccinated<br>(n =37,402) | 16 | 77.2 [59.5-86.3] | 80.0 [64.3-88.0] |
|  | Vaccinated<br>(n = 809,481) | 79 |  |  |
| Comorbidities<br>(n =266,097) | Unvaccinated<br>(n =9,590) | 14 | 85.1 [72.1-91.4] | 81.8 [65.6-89.7] |
|  | Vaccinated<br>(n = 256,507) | 56 |  |  |
| No comorbidities<br>(n =1,270,338) | Unvaccinated<br>(n = 86,456) | 30 | 76.2 [63.5-83.9] | 75.7 [62.7-83.7] |
|  | Vaccinated<br>(n = 1,183,882) | 98 |  |  |
| Male<br>(n =769,170) | Unvaccinated<br>(n = 51,459) | 19 | 69.1 [47.5-80.8] | 69.6 [48.1-81.3] |
|  | Vaccinated<br>(n =717,711) | 82 |  |  |
| Female<br>(n = 767,265) | Unvaccinated<br>(n = 44,587) | 25 | 82.2 [71.5-88.6] | 84.1 [74.2-89.8] |
|  | Vaccinated<br>(n = 722,678) | 72 |  |  |
| Infection-Naive<br>(n =1,534,846) | Unvaccinated<br>(n = 95,926) | 42 | 70.6 [58.1-79.0] | 68.3 [54.6-77.5] |
|  | Vaccinated<br>(n =1,438,920) | 150 |  |  |

Effectiveness is expressed as % [CI<sub>95%</sub>]
\*Adjusted or stratified for age group, gender and presence of comorbid conditions.

**Figure 4.**
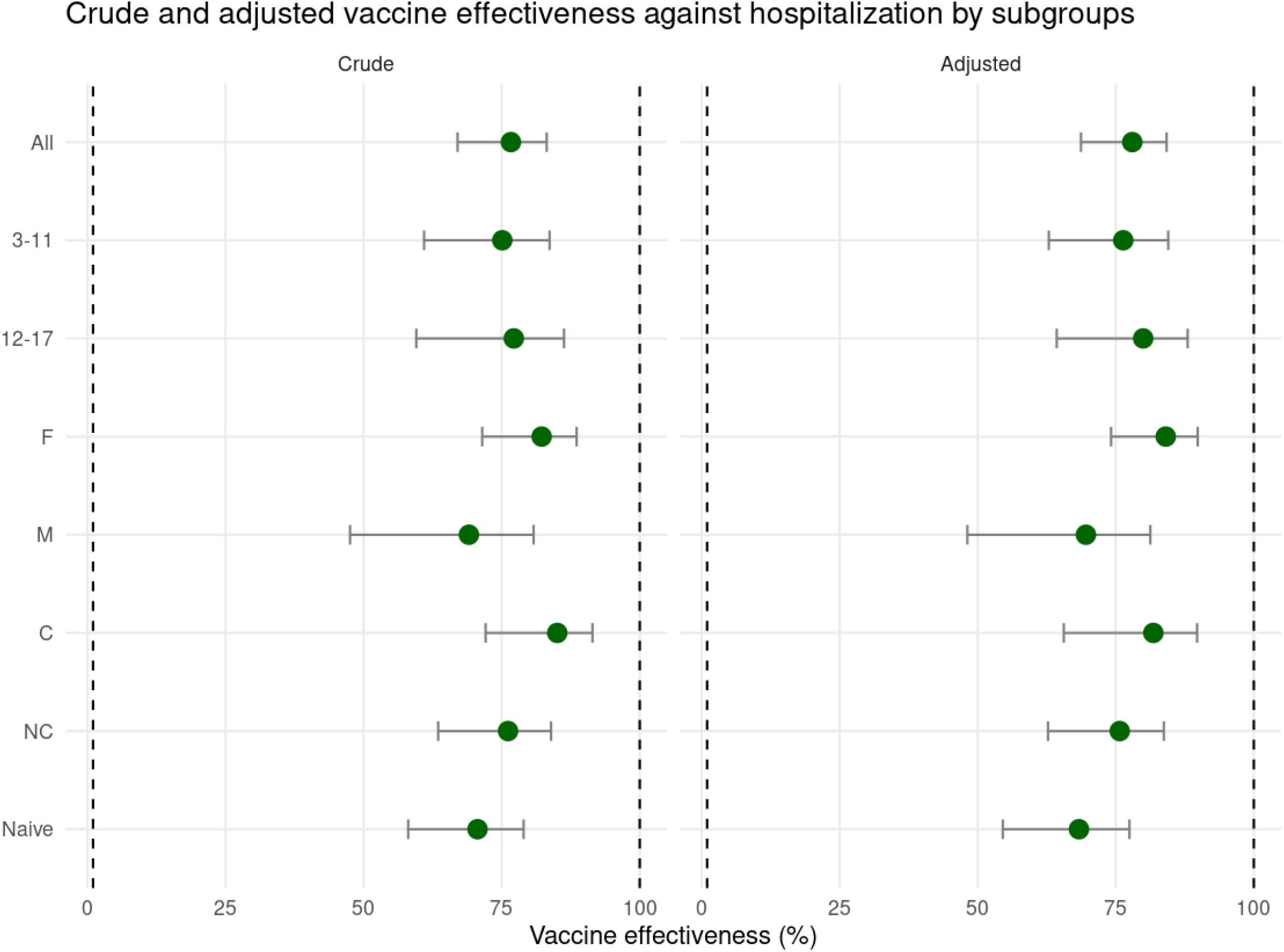
Vaccine effectiveness for different categories: 3-11 and 12-17 age subgroups, Females (F), Males (M), subjects with comorbidities (C) and without them (NC), and SARS-CoV-2 infection naive-subjects (Naive). Error bars indicate the %[CI95%].

When subjects with previously documented SARS -CoV-2 infections were excluded, vaccine effectiveness was 68·3 [54·6-77·5] (adjusted) and 70·6 [58·1-79·0] (crude), comparable to results in the primary analysis. (Table 3 and Figure 4).

Crude and adjusted VE for the entire population and age subgroups decreased in the entire population during the Omicron period compared to the previous period. (Table 4). When VE was analysed by age subgroup, it decreased in the 3-11-year subgroup but remained similar in the adolescents (Table 5).

**Table 4:** Vaccine effectiveness by study period in the entire population

| Study period | Unvaccinated<br>(n events) | Vaccinated<br>(n events) | Crude<br>effectiveness<br>%, [CI95%] | Adjusted<br>effectiveness*<br>%, [CI95%] |
| --- | --- | --- | --- | --- |
| 14 December<br>2021- 18<br>January 2022 | 30 | 84 | 81.3<br>[71.7-87.7] | 82.7<br>[73.2-88.6] |
| 19 January - 9<br>March 2022 | 14 | 70 | 66.6<br>[40.8-81.2] | 67.7<br>[39.9-81.4] |

Effectiveness is expressed as % [CI<sub>95%</sub>]
\*Adjusted for age group, gender, and presence of comorbid conditions.

**Table 5:**
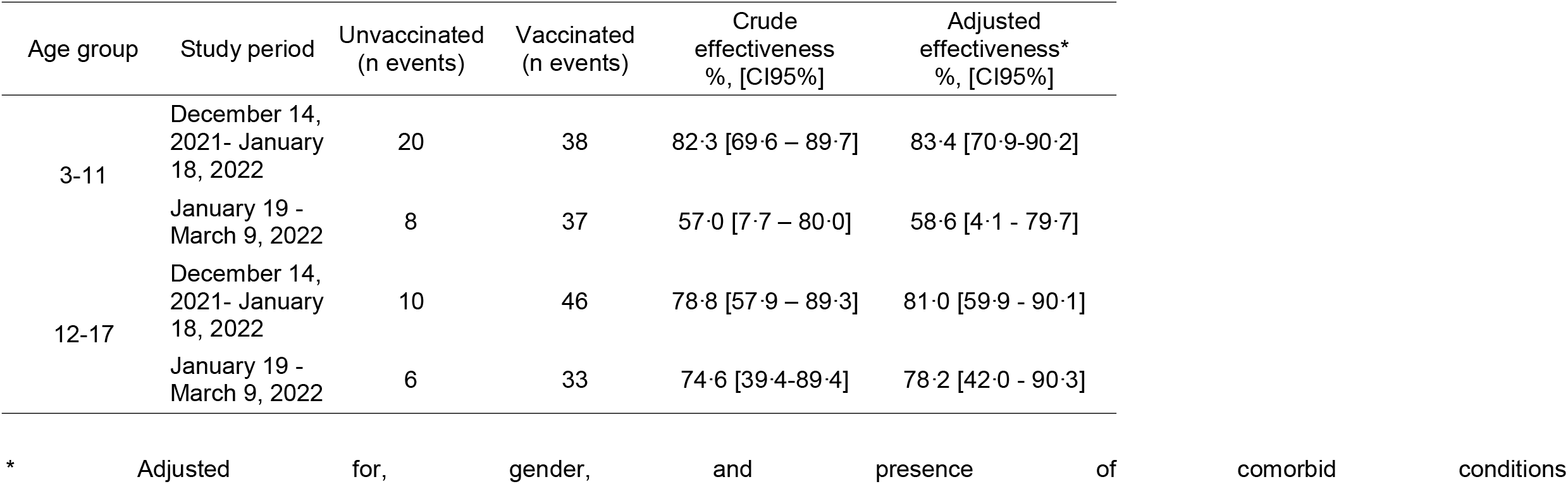
Vaccine effectiveness by study period and age group.

| Age group | Study period | Unvaccinated<br>(n events) | Vaccinated<br>(n events) | Crude<br>effectiveness<br>%, [CI95%] | Adjusted<br>effectiveness*<br>%, [CI95%] |
| --- | --- | --- | --- | --- | --- |
| 3-11 | December 14,<br>2021- January<br>18, 2022 | 20 | 38 | 82.3 [69.6 – 89.7] | 83.4 [70.9-90.2] |
|  | January 19 -<br>March 9, 2022 | 8 | 37 | 57.0 [7.7 – 80.0] | 58.6 [4.1 - 79.7] |
| 12-17 | December 14,<br>2021- January<br>18, 2022 | 10 | 46 | 78.8 [57.9 – 89.3] | 81.0 [59.9 - 90.1] |
|  | January 19 -<br>March 9, 2022 | 6 | 33 | 74.6 [39.4-89.4] | 78.2 [42.0 - 90.3] |

\* Adjusted for, gender, and presence of comorbid conditions

## DISCUSSION

This study in real-world settings demonstrates that paediatric vaccination, 14 days or more from two-dose-scheme, prevented 78·0% of COVID-19-associated hospitalisations during the Omicron outbreak in the Province of Buenos Aires. VE for the 3-11 and 12-17 age subgroups was 76·4% and 80·0%, respectively. VE for the entire population was 82·7% during the period of Delta and Omicron overlapping circulation and decreased to 67·7% when Omicron was the only variant present. These results are consistent with recent studies conducted in similar age groups. ^24-27^

VE against hospitalisation, ICU admission and death in vaccine clinical trials and in subsequent real-world studies, including previous variants of SARS-COV-2 in the paediatric population, was higher than 90%. ^14-17,22,23^ However, since the identification of Omicron VOC, new data has become available reporting a significant reduction of the immune sera neutralising capacity of vaccinated or infected subjects with this variant compared to previous ones.^2,3^ Surveillance reports in the adult population from the UK during the Omicron breakout indicate that after a two-dose-scheme, VE against hospitalisation plummeted from 65-85% within 3 months of vaccination to 30-35% after 6 months. By contrast, during Delta’s predominance, VE against hospitalisations fell from 95-99% to 70-85% in a similar period.^6^

With respect to the paediatric population, the Centres for Disease Control and Prevention provided the earliest data about VE in 5-11-year-old children carried out in real-world settings during the Omicron period. They reported a VE against hospitalisation of 74% [–35 - 95] with wide confidence intervals that included zero with two doses of BNT162b2; this figure is near the range of our data which was 76·4% [62·9-84·5] over the entire study period.^24^ However, when we analysed VE in the 3-11-year subgroup during the Omicron predominance period, a marked decrease was evident. A preprint study carried out in New York from 29 November 2021 to 30 January 2022 reported that VE against hospitalisation of a two-dose scheme of BNT162b2 was 48% for the 5-11-age subgroup in the last week of the study.^25^ In a recent case-control test-negative study carried out in the US, VE against hospitalisation was 68% for the 5-11-year subgroup.^26^ A recent Chilean study performed in 3-5-year-old children who received a two-dose scheme of an inactivated vaccine (CoronaVac) during the Omicron outbreak reported a VE against hospitalisation and ICU admission of 64% and 69%, respectively. Our findings are consistent with published studies.^27^ Researchers, in general, attributed the early decrease of VE to the ability to evade immune response of Omicron, to waning immunity, or to both; and this might signal the potential need to review vaccination schemes and/or dosing.^25-27^ To our knowledge, ours is the first study carried out in a real-world setting for the 3-11 age population receiving the full scheme of BBIBP-CorV and reporting VE against hospitalisation.

Concerning the adolescent population, the CDC reported high VE for the two-dose scheme of BNT162b2. Within 5 months of full vaccination, VE was 92% and 94%, for the 12–15 and 16–17 age groups, respectively; however, it decreased after 5 months to 73% and 88%, for the same age subgroups during a period of Delta and Omicron circulation. Furthermore, more recent studies have shown even lower VE. ^24,25^ Data from the New York study conducted from 29 November 2021 to 30 January 2022 indicated that VE fell from 94% to 73% in 12-17-year-old patients with two-doses of BNT162b2.^25^ Moreover, in a recent case-control test-negative study carried out in the US, VE against hospitalisation was 40% [9-60], which corresponds to VE of 79% [51-91] against critical COVID-19 and 20% [-25-49] against noncritical COVID-19. The authors suggested that the lower VE might have been associated with the amount of time since vaccination and the emergence of Omicron.^26^ We found a VE of 80·0% [64·3-88·0] against hospitalisation during the entire study period, but 81·0% and 78·2% for the two periods of different VOCs circulation. These figures are within the range of the abovementioned studies.^24-27^

The different fall of VE between children and adolescents over the two periods was noticeable, in line with recent research that has also suggested that VE might be lower in younger children.^25,27^ This might be ascribed to a less robust immunological response in this population.^29^ Therefore, they might need higher doses than the administered ones, different intervals between them, additional shots, or the utilisation of vaccines designed to target different antigens.^25^

In the adult population, waning VE against hospitalisation is reversed with the administration of a booster dose.^4,5^ Furthermore, VE against COVID-19 symptomatic disease improved in adolescents with a booster dose, as reported by the CDC.^24^

Argentina started the administration of a booster dose in November 2021, prioritising high-risk population groups, followed by vaccination of the adolescent group. With respect to the 3-11 subpopulation, neither the National Immunisation Commission nor the WHO recommend the administration of a booster dose to this age subgroup.^28^ However, more severe clinical presentations in the paediatric population have been reported due to Omicron BA.1 and BA.2 sub-lineages.^10-12^ Two recent studies from South Africa and Hong Kong performed during the Omicron wave recorded more severe disease in children, including laryngeal obstruction and neurological symptoms, with increased requirement of ICU admission.^10,11^ Further research is needed to assess the need for a booster dose in 3-11-year-old children.

### Limitations

The first limitation of this study lies in its observational nature. Therefore, If the vaccinated and unvaccinated groups are systematically different, VE might be affected. We addressed this problem by adjusting our risk estimation with the available confounders. However, residual confounders cannot be discarded. Second, in the 3-11-age subgroup we could only analyse VE for the BBIBP-CorV vaccine as it was the only vaccine available in Argentina. Third, we only were able to estimate VE in adolescents for the two vaccines of the mRNA vaccine platform without differentiating between them, due to the scarcity of events. Fourth, we could only assess VE of a two-dose schedule vaccination for a median time since vaccination of 114 days. Nevertheless, this limitation is akin to most studies conducted in children, given the recent start of the vaccination rollout. A longer follow-up period after vaccination will be crucial to evaluate the duration of the effectiveness against COVID-19– associated hospitalisation and death. Fifth, a certain degree of misclassification cannot be ruled out, especially concerning potential contamination by incidental COVID-19 cases. To deal with this issue, we considered only those hospitalisations occurring within 14 days from COVID-19 diagnosis. Sixth, we could not assess the effect of a booster dose on VE in adolescents, given that boosters were only recently authorised in this subgroup in February 2022. Seventh, notwithstanding the high incidence of Omicron cases, there were still relatively few child-hospital admissions, so we were not able to estimate markers of severity of disease, such as ICU admission or death. Eighth, genetic characterization of patients’ viruses was not available, therefore Delta and Omicron predominance periods were based on surveillance data. Ninth, the estimate of VE corresponds to Omicron.BA.1, the lineage of Omicron circulating during the study period; it is possible that VE could change if new SARS-CoV-2 variants emerge. Tenth, we could not assess a possible waning effect due to the recent vaccination of the 3-11-age subgroup and the scarcity of events. Finally, this study includes an adolescent group and 3-11-year-old children; these subpopulations might differ in the exposures, or in the test-seeking behaviours. ^25^

## Conclusions

This report provides real-world evidence of vaccine protection against COVID-19-associated hospitalisations among children and adolescents aged 3–17 years during the Omicron outbreak; VE effect was consistent in all subgroups tested. Nevertheless, VE decreased in the period of Omicron predominance, especially in children aged 3-11. Application of a booster dose in this subpopulation warrants further consideration.

## Data Availability

All data produced in the present study are available upon reasonable request to the authors.

## Notes

### Author Declarations

The Central Ethics Committee of the Ministry of Health of the Province of Buenos Aires evaluated and approved the protocol of the present study on 18 March 2022. The report number ACTA-2022-07714920-GDEBA-CECMSALGP. Informed Consent This study was exempted of informed consent due to its retrospective nature, and given it is a public health-related official programme. Anonymization of data Data were anonymized by the following procedure: The personal ID number was used to link the databases of follow-up and vaccination. After this process, we removed the personal ID number and created an ID reference number for each individual. This reference number is not associated with any personal information.

## REFERENCES

[1] Liu Q, Qin C, Liu M, Liu J. Effectiveness and safety of SARS-CoV-2 vaccine in real-world studies: a systematic review and meta-analysis. Infect Dis Poverty. 2021;10(1):132. doi: 10.1186/s40249-021-00915-3. PMID: 34776011;

[2] Cele S, Jackson L, Khoury DS, et al. Omicron extensively but incompletely escapes Pfizer BNT162b2 neutralization. Nature 2022; 602(7898): 654–656.

[3] VanBlargan, L.A., Errico, J.M., Halfmann, P.J. et al. An infectious SARS-CoV-2 B.1.1.529 Omicron virus escapes neutralization by therapeutic monoclonal antibodies. Nat Med 2022; 28:490–495

[4] Abu-Raddad LJ, Chemaitelly H, Ayoub HH, et al. Effect of mRNA Vaccine Boosters against SARS-CoV-2 Omicron Infection in Qatar. N Engl J Med. 2022 Mar 9:NEJMoa2200797. doi: 10.1056/NEJMoa2200797. [Epub ahead of print].

[5] Andrews N, Stowe J, Kirsebom F, et al. Covid-19 Vaccine Effectiveness against the Omicron (B.1.1.529) Variant. N Engl J Med. 2022 Mar 2:NEJMoa2119451. doi: 10.1056/NEJMoa2119451. [Epub ahead of print].

[6] UK Health Security Agency. COVID-19 vaccine surveillance report – week 11. [Internet] 2022. [cited 2022 Mar 18]. Available from: https://t.co/1cGniY6AgW

[7] Ministerio de Salud Argentina. Informes de vigilancia genómica. [Internet] 2022 Jan. [cited 2022 Mar 26]. Available from: https://www.argentina.gob.ar/salud/coronavirus-COVID-19/informacion-epidemiologica/enero-2022-0

[8] Plotkin SA and Levy O. Considering Mandatory Vaccination of Children for COVID-19. Pediatrics 2021;147(6):e2021050531

[9] Waon severity in children under 5 years old before and after Omicron emergence in the US. [Preprint 2022 Jan 13]. NIH. Available from: DOI: 10.1101/2022.01.12.22269179

[10] Cloete J, Kruger A, Masha M, et al. Paediatric hospitalisations due to COVID-19 during the first SARSCoV-2 omicron (B.1.1.529) variant wave in South Africa: a multicentre observational study. Lancet Child Adolesc Health [Internet] 2022, Feb 18 [cited 2022 Mar 21]:1–9. Available from: https://doi.org/10.1016/S2352-4642(22)00027-X

[11] Tso W, Kwan M, Wang Y.L. et al. Intrinsic severity of SARS-CoV-2 Omicron BA.2 in uninfected, unvaccinated children: a population-based, case-control study on hospital complications. [Preprint, 2022 Mar 21]. The Lancet. Available from: https://papers.ssrn.com/sol3/papers.cfm?abstract_id=4063036.

[12] Marks KJ, Whitaker M, Agathis NT, et al. Hospitalization of Infants and Children Aged 0–4 Years with Laboratory-Confirmed COVID-19 — COVID-NET, 14 States, March 2020– February 2022. MMWR Morb Mortal Wkly Rep. 2022 Mar 18;71(11):429–436

[13] Xia S, Zhang Y, Wang Y, et al. Safety and immunogenicity of an inactivated COVID-19 vaccine, BBIBP-CorV, in people younger than 18 years: a randomised, double-blind, controlled, phase 1/2 trial. Lancet Infect Dis. 2022 Feb;22(2):196–208.

[14] Walter EB, Talaat KR, Sabharwal C, et al. Evaluation of the BNT162b2 Covid-19 vaccine in children 5 to 11 years of age. N Engl J Med 2022;386:35–46.

[15] Frenck RW Jr, Klein NP, Kitchin N, et al.; C4591001 Clinical Trial Group. Safety, immunogenicity, and efficacy of the BNT162b2 Covid-19 vaccine in adolescents. N Engl J Med 2021;385:239–50.

[16] Polack FP, Thomas SJ, Kitchin N, et al.; C4591001 Clinical Trial Group. Safety and efficacy of the BNT162b2 mRNA Covid-19 vaccine. N Engl J Med 2020;383:2603–15.

[17] Ali K, Berman G, Zhou H, et al. Evaluation of mRNA-1273 SARS-CoV-2 vaccine in adolescents. N Engl J Med 2021; 385:2241–2251.

[18] Ministerio de Salud Argentina. Actualización de los Lineamientos Técnicos Resumen de recomendaciones vigentes para la Campaña Nacional de Vacunación contra la COVID-19. [Internet] 2022. [cited 2022 Mar 18] Available from: https://bancos.salud.gob.ar/recurso/actualizacion-de-los-lineamientos-tecnicos-resumen-de-recomendaciones-vigentes-para-la

[19] Olson SM, Newhams MM, Halasa NB, et al. Effectiveness of BNT162b2 Vaccine against Critical Covid-19 in Adolescents. N Engl J Med. 2022;386(8):713–723.

[20] Zambrano LD, Newhams MM, Olson SM, et al. Effectiveness of BNT162b2 (Pfizer-BioNTech) mRNA Vaccination Against Multisystem Inflammatory Syndrome in Children Among Persons Aged 12–18 Years — United States, July–December 2021. MMWR Morb Mortal Wkly Rep 2022;71:52–58.

[21] Levy M, Recher M, Hubert H, et al. Multisystem inflammatory syndrome in children by COVID-19 vaccination status of adolescents in France. JAMA 2022;327(3):281–283.

[22] Jara A, Undurraga E, Flores J, et al. Effectiveness of an inactivated SARS-CoV-2 vaccine in children and adolescents: A large scale observational study. [Preprint, 2022 Feb 15]. The Lancet. Available from: https://papers.ssrn.com/sol3/papers.cfm?abstract_id=4035405

[23] Glatman-Freedman A, Hershkovitz Y, Kaufman Z, et al. Effectiveness of BNT162b2 Vaccine in Adolescents during Outbreak of SARS-CoV-2 Delta Variant Infection, Israel, 2021. Emerging Infectious Diseases 2021; 27(11):2919–22.

[24] Klein NP, Stockwell MS, Demarco M, et al. Effectiveness of COVID-19 Pfizer-BioNTech BNT162b2 mRNA Vaccination in Preventing COVID-19–Associated Emergency Department and Urgent Care Encounters and Hospitalizations Among Nonimmunocompromised Children and Adolescents Aged 5–17 Years — VISION Network, 10 States, April 2021–January 2022. MMWR Morb Mortal Wkly Rep 2022;71:352–358.

[25] Dorabawila V, Hoefer D, Bauer U, et al. Effectiveness of the BNT162b2 vaccine among children 5-11 and 12-17 years in New York after the Emergence of the Omicron Variant. [Preprint, 2022 Feb 28]. Available from: https://www.medrxiv.org/content/10.1101/2022.02.25.22271454v1.full.pdf

[26] Price AM, Olson SM, Newhams MM, et al. BNT162b2 Protection against the Omicron Variant in Children and Adolescents. N Engl J Med. 2022 Mar 30. doi: 10.1056/NEJMoa2202826. [Epub ahead of print].

[27] Araos R, Jara A, Undurraga E, et al. Effectiveness of CoronaVac in children 3 to 5 years during the omicron SARS-CoV-2 outbreak. [Preprint, 2022 Mar 15]. Nature portfolio. Available from: https://www.researchsquare.com/article/rs-1440357/v1

[28] WHO SAGE roadmap for prioritizing use of COVID-19 vaccines 2022. Available from: https://www.who.int/groups/strategic-advisory-group-of-experts-on-immunization/covid-19-materials (accessed April 4, 2022).

[29] Semmes EC, Chen JL, Goswami R, Burt TD, Permar SR, Fouda GG. Understanding Early-Life Adaptive Immunity to Guide Interventions for Pediatric Health. Front Immunol. 2021;11:595297. Published 2021 Jan 21. doi:10.3389/fimmu.2020.595297

